# Are epidemic growth rates more informative than reproduction numbers?

**DOI:** 10.1101/2021.04.15.21255565

**Authors:** Kris V Parag, Robin N Thompson, Christl A Donnelly

## Abstract

Summary statistics, often derived from simplified models of epidemic spread, inform public health policy in real time. The instantaneous reproduction number, *R*_*t*_, is predominant among these statistics, measuring the average ability of an infection to multiply. However, *R*_*t*_ encodes no temporal information and is sensitive to modelling assumptions. Consequently, some have proposed the epidemic growth rate, *r*_*t*_, i.e., the rate of change of the log-transformed case incidence, as a more temporally meaningful and model-agnostic policy guide. We examine this assertion, identifying if and when estimates of *r*_*t*_ are more informative than those of *R*_*t*_. We assess their relative strengths both for learning about pathogen transmission mechanisms and for guiding public health interventions in real time.

## 1 INTRODUCTION

Inferring changes in pathogen transmissibility during epidemics is an important challenge. Increases in transmissibility may forewarn of elevating caseloads and hospitalisations, while decreasing rates of spread may evidence the effectiveness of earlier interventions or the influence of infection-acquired immunity (Anderson and May, 1991). Practical limitations on the scope and speed of outbreak surveillance mean that population-wide summary statistics, derived from simplified models of epidemics, often inform public health policy in real time. The instantaneous reproduction number at time *t*, denoted *R*_*t*_, is predominant among these statistics. It measures the average number of secondary infections generated per effective primary case at that time. An *R*_*t*_ above (or below) 1 indicates a growing (or waning) epidemic and often forms part of the evidence base for policy decisions on the imposition or release of interventions (Anderson et al., 2020). However, *R*_*t*_ encodes no temporal information. For example, *R*_*t*_ = 2 indicates approximate epidemic doubling (per generation of infections) but not the speed of that doubling. Moreover, because inference of *R*_*t*_ depends on the model used (and hence its assumptions), differing estimates may be obtained from the same data, complicating the interpretation of *R*_*t*_ as a signal for epidemic response (Lloyd, 2009; Parag and Donnelly, 2020).

Consequently, the instantaneous epidemic growth rate, *r*_*t*_, defined as the rate of change of the log-transformed case incidence, has been proposed as a more informative and understandable measure of transmission dynamics (Pellis et al., 2020). Growth rates may be estimated directly from the gradient of the log-transformed observed incidence curve, have a natural temporal interpretation as the speed of case accumulation and still encode key dynamics e.g., the sign of *r*_*t*_ and *R*_*t*_ − 1 signify similar transmission trends. Estimates of *r*_*t*_ can therefore, seemingly, be derived independently of an epidemic model. However, if a model is assumed, there is a one-to-one correspondence between *r_t_* and *R*_*t*_. Thus *R_t_* may provide no more information about transmission patterns than that available already from *r*_*t*_ (Wallinga and Lipsitch, 2007). While these observations may at first recommend *r*_*t*_ as the more useful measure for policymaking, there are implicit complications.

First, when comparing transmission across different spatial scales, epidemic phases or even data types (e.g., hospitalisations or cases), a non-dimensional parameter may be more useful. A value of *R*_*t*_ = 2 has the same interpretation of a primary case generating two secondary ones on average, regardless of the region studied or the phase of the epidemic considered, with important implications for interventions (e.g. if *R*_*t*_ = 2, then more than half of transmissions must be prevented for the epidemic to start declining). Second, the process of estimating the logarithmic derivative of a noisy incidence curve is not trivial and noise-smoothing choices may actually be equivalent to modelling assumptions. Third, information encoded in *R*_*t*_ may be more easily leveraged to develop other useful outbreak analytics, such as probabilities of epidemic elimination (Parag et al., 2020) or herd immunity thresholds (see Discussion and Hethcote (2000)). Last, biases and delays in reporting and surveillance may have differing impacts on estimates of both quantities, making it unclear which offers the higher fidelity view of transmission (Lloyd, 2009).

In this paper, we investigate and discuss the various complexities and subtleties mediating the practical informativeness of estimates of both *R*_*t*_ and *r*_*t*_, which we denote 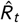 and 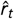, respectively. We outline how these quantities can be computed from incidence curves using renewal models and smoothing filters. This leads us to our main result: that the smoothing assumptions inherent in obtaining 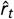 from noisy incidence curves can be in some senses equivalent to the epidemiological ones necessary for obtaining 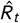. Consequently, we conclude that the question of whether *R*_*t*_ or *r*_*t*_ is more informative for real-time public health policymaking depends on the relative accuracy of the epidemiological assumptions and on how well the subtleties and uncertainties underlying each summary statistic are communicated. Estimates of *R*_*t*_ and *r*_*t*_ in combination, alongside contextual information about the ongoing epidemic, will provide the most complete picture of pathogen transmission and control.

## 2 METHODS

### 2.1 Computing reproduction numbers and growth rates

Inferring the time-varying transmissibility of a pathogen from routinely available surveillance data is vital to assess ongoing and upcoming trends in epidemic dynamics. Among the most common data types is the incidence curve, which represents the time-series of of new cases. We use *I*_*t*_ to denote the incidence at time *t* and let *w*_*j*_ be the probability that a primary case takes *j* time units (usually in days) to generate a secondary case. The set of *w*_*j*_ for all *j* constitutes the generation time distribution of the disease, where we make the common assumption that the generation time distribution is approximated by the serial interval distribution (Wallinga and Teunis, 2004; Cori et al., 2013). The serial interval distribution describes the times between symptom onsets for primary and secondary cases and is often computed from independent line-list data (Cowling et al., 2009; Hart et al., 2021). We assume that the set of *w*_*j*_ has been well characterised for the infectious disease of interest.

The renewal model (Fraser, 2007) relates the instantaneous reproduction number at time *t, R*_*t*_, to the incidence curve and generation time distribution as in Eq. (1) with 𝔼 [*I*_*t*_] indicating the mean of *I*_*t*_. Typically, an assumed distribution (e.g. Poisson or negative binomial) is used to statistically relate this mean to *I*_*t*_, and estimates of *R*_*t*_ (i.e. 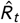) are obtained using various Bayesian or maximum likelihood computational approaches.

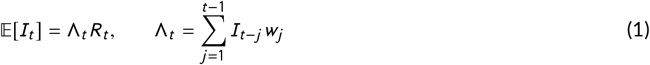

The total infectiousness, Λ_*t*_, summarises how past incidence propagates forwards in time by incorporating knowledge of the generation time distribution via a convolution. Many approaches exist for inferring 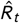 from the incidence curve {*I*_1_, *I*_2_, …, *I*_*T*_} with *T* as the last observed time (see (Anderson et al., 2020) for more details). These estimates, 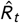, are increasingly employed for tracking transmissibility during epidemics and guiding public health responses.

The instantaneous growth rate, *r*_*t*_, has been used less frequently to assess transmissibility over time but has recently gained attention as an alternative to *R*_*t*_ (Pellis et al., 2020; Dushoff and Park, 2021) and is among the metrics that COVID-19 advisory bodies track. The quantity 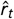 can be derived from {*I*_1_, *I*_2_, …, *I*_*T*_} without additional epidemiological knowledge or assumptions e.g., no estimated generation time distribution is required. Instead, the logarithmic derivative of some smoothed version of the incidence, 𝕊 [*I*_*t*_], is used, as in the left side of Eq. (2).

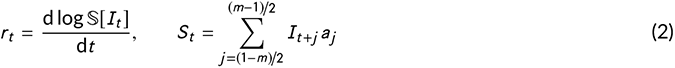

Here 𝕊[*I*_*t*_] refers to any smoothing, which at time *t* may depend on any subset of the incidence curve (and not just *I*_*t*_). There are various ways of deriving 𝕊 [*I*_*t*_] curves (e.g. using splines or moving average filters (Pellis et al., 2020)).

We can unify many of these approaches to smoothing within the framework of Savitzky-Golay (SG) filters (Savitzky and Golay, 1964). SG filters, with dimension *m* and coefficients *a*_*j*_, perform local least-squares polynomial smoothing via the discrete convolution or kernel in the right side of Eq. (2). We denote the resulting smoothed incidence as 𝕊 [*I*_*t*_] = *S*_*t*_ and we may optimise the *a*_*j*_ coefficients via least squares or select their values to confer some desired properties (for example to maintain certain waveform or frequency characteristics of the original data). We can realise a standard moving average filter within the SG framework by setting each *a*_*j*_ = 1/*m*, for example. The SG framework has broad applications and further information on its uses and properties can be found in (Schafer, 2011).

The reproduction numbers and growth rates we consider should not be confused with the basic reproduction number, *R*_0_, and the intrinsic growth rate, *r*, which can be estimated using numerous methods (e.g., via compartmental or Richards’ growth models (Yan and Chowell, 2019)) from various data sources (e.g., prevalence or cumulative case data). While these are related to our *R*_*t*_ and *r*_*t*_ during the earliest phases of an epidemic, *R*_0_ and *r* cannot track time-varying changes in transmissibility. Instead they provide insight into initial epidemic growth upon invasion (Anderson et al., 2020). The methods above do not consider spatial, contact or other heterogeneities. However, including these may not always improve transmissibility estimates (Liu et al., 2018). Next, we clarify how *R*_*t*_ and *r*_*t*_ are related.

### 2.2 Connecting reproduction numbers and growth rates

Eq. (1) and Eq. (2) describe simple and general approaches to estimating *R*_*t*_ and *r*_*t*_ from an incidence curve. While a known generation time distribution or serial interval is assumed in Eq. (1) when determining 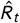, Eq. (2) neither makes mechanistic assumptions nor requires additional data for calculating 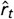. However, if the assumptions in Eq. (1) are made then it is possible to derive a model-dependent 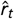 from 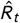. The generalised method for connecting these two summary statistics is given in the left side of Eq. (3) (Wallinga and Lipsitch, 2007), with 𝕄_*w*_ denoting the moment generating function about the generation time distribution (defined by the set of *w*_*j*_ from Eq. (1)).

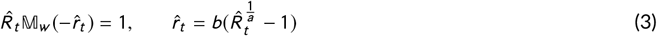

The left side of Eq. (3) states that the relationship between 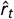 and 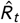 depends strongly on the parametric form of the generation time (or in practice, the serial interval) distribution. The set of *w*_*j*_ is most commonly parametrised from the gamma family of distributions with shape and scale parameters *a* and *b*. This leads to the analytic expression on the right side of Eq. (3). Although the moment generating approach suggests a general way of connecting 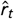 and 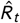, there is an implicit exponential growth or decay assumption within this formula (Wallinga and Lipsitch, 2007). While Eq. (3) is developed for a general renewal model framework, we can also specialise this method to popular compartmental models. For example, under a linearised compartmental Susceptible-Infectious-Recovered (SIR) model, we obtain 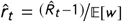, with 𝔼 [*w*] as the mean generation time (Bettencourt and Ribeiro, 2008).

## 3 RESULTS

We examine how the model-based 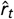 relates to 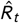 under a given generation time distribution (see Methods). The gamma and SIR simplifications of Eq. (3) provide key insights into the relative informativeness of these statistics. First, we see that the sign of 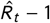 and 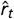 are equivalent, making either equally good for inferring the transitions between growing and declining epidemics. We illustrate this for a simulated epidemic in Fig. 1, which has been constructed to model seasonal transmission dynamics. The example we provide is representative of the range of possible epidemic trajectories (and estimates) that would result from our chosen true sinusoidal *R*_*t*_ profile.

**FIGURE 1.**
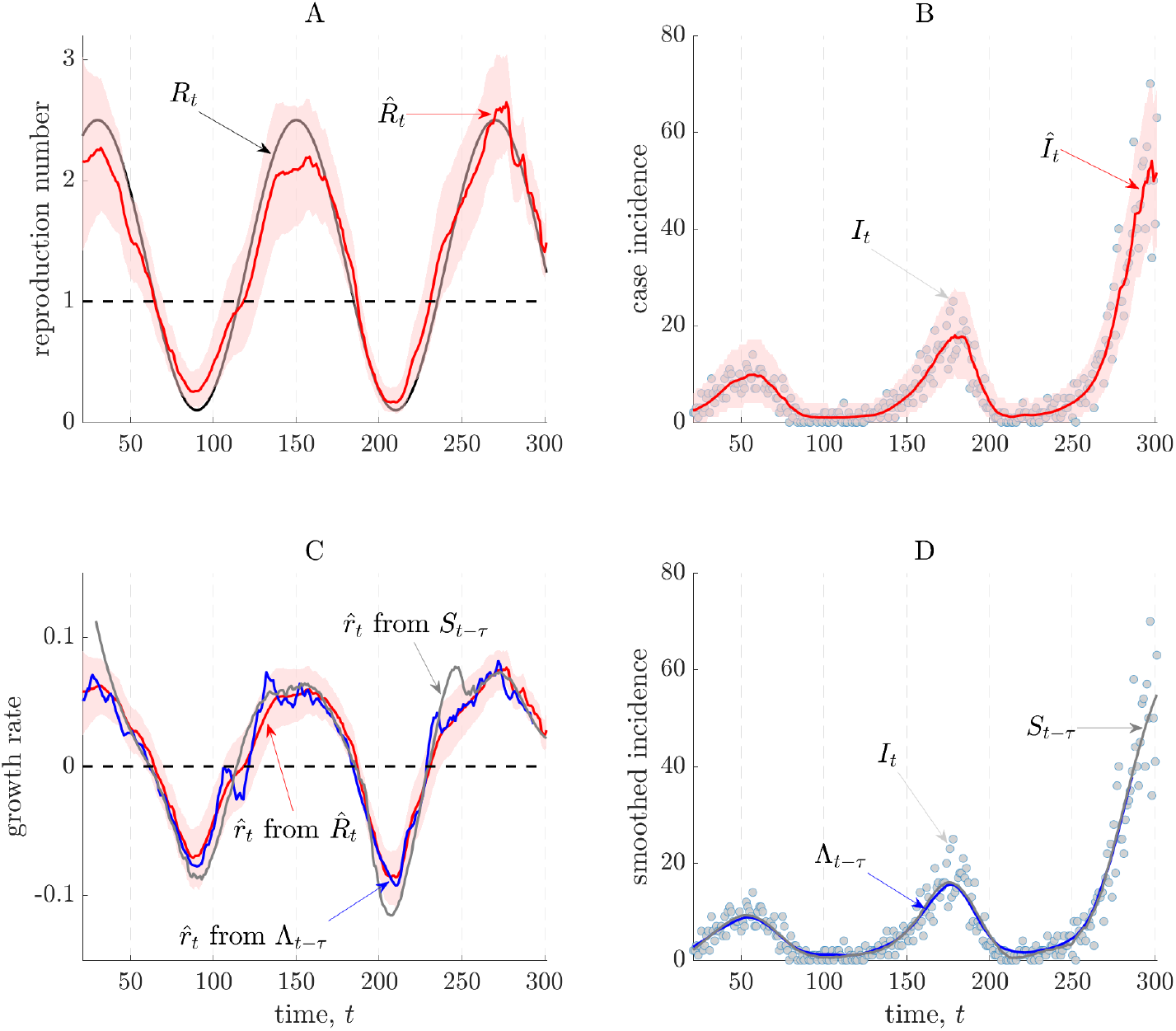
Instantaneous reproduction numbers and growth rates. We simulate a seasonally varying epidemic with incidence *I*_*t*_, according to the renewal model with true transmissibility *R*_*t*_ and serial interval distribution estimated for Ebola virus from (Van Kerkhove et al., 2015). In panels A and B, we estimate the instantaneous reproduction number 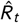 (with 95% credible intervals) using EpiFilter (see (Parag, 2020)) and provide one-step-ahead predictions 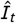 using 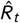. In panels C and D we derive three growth rate estimates, 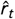 using: 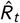 (via the (Wallinga and Lipsitch, 2007) approach), a smoothed-shifted version of the incidence curve *S*_*t* \**τ*_ (via SG filters) and a shifted version of the total infectiousness of the epidemic Λ_*t* \**τ*_ by treating it as a type of SG filter.

We compute 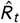 using the EpiFilter method (Parag et al., 2020) (red, A), which provides minimum mean squared error Bayesian estimates. We assume knowledge of the true generation time distribution and validate our estimates with one-step-ahead incidence predictions (red, B) as in (Parag and Donnelly, 2020). The intersections of 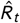 (red, A) with 1 and those of the model-based 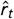 (red, C) with 0 coincide, as expected from Eq. (3). Both provide consistent assessments of time-varying transmission, correctly signalling rising and falling seasons.

We next compute the model-agnostic, log-derivative-based 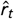 using an SG filter as in Eq. (2), which effectively fits local splines to the incidence curve. This estimate (grey, C) correlates well with our model-based one (red, C), with some overshoot in periods where incidence is small (and estimation known to be more difficult (Parag et al., 2020)). The only assumptions made in obtaining this estimate relate to how we smooth the data to obtain stable log-derivatives (e.g., we have to make choices about the order of our splines or the dimension of our moving filters). Current approaches to deriving model-agnostic 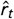 values must all ultimately make similar assumptions and choices (Pellis et al., 2020). Having made these key observations, our main result emerges.

Comparing Eq. (1) and Eq. (2), we see that the total infectiousness, Λ_*t*_, is an implicit SG filter, with the value of *m* determined by the support of the generation time distribution. Hence, we construct another growth rate estimate, 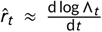 as shown in Fig. 1 (blue, C). This estimate matches the other two growth rate estimates well but with a decreased overshoot. This correspondence is novel and, importantly, clarifies how time-varying model-agnostic growth rates and instantaneous reproduction numbers relate by exposing that the generation time distribution is effectively an epidemiologically informed smoothing filter.

We confirm this by comparing Λ_*t*_ and *S*_*t*_ (grey and blue, D), which are effectively two possible realisations of 𝕊 [*I*_*t*_] from Eq. (2). These are written Λ_*t* \**τ*_ and *S*_*t* \**τ*_ to indicate that they have been shifted to remove lags, *τ*, which naturally result from applying smoothing filters. Generally, *τ* is related to the mean generation time. We do not provide credible intervals for the two SG-based 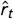 here as we simply intend these results to demonstrate proof-of-concept. Note that all Bayesian credible intervals we present are equal tailed (based on relevant quantiles).

Last, in Fig. 2 we examine robustness to misspecification of the generation time distribution. We use the same inference procedures as above but now the mean generation time assumed in estimation has a mean that is 33% smaller than that of the true Ebola virus distribution (under which the data is generated). Misspecification could occur due to interventions or other epidemiological changes (Ali et al., 2020). We find that 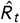 is sensitive to this change (compare the red and blue estimates in A). However, the 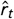 computed from the misspecified 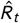 is mostly stable, although there is increased uncertainty (compare the red and blue estimates in B). Correspondence between model-based and model agnostic 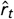 is also maintained but not shown.

**FIGURE 2.**
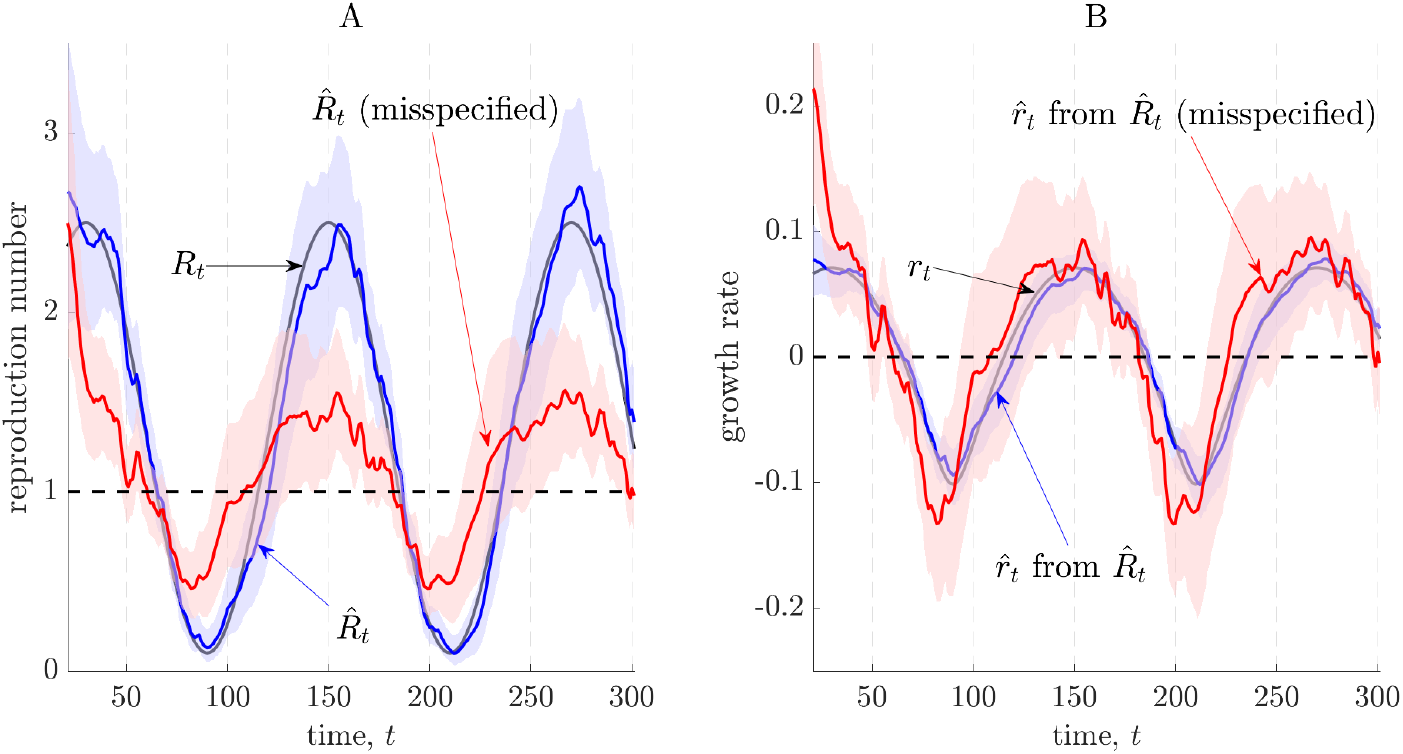
Misspecified estimates of reproduction numbers and growth rates. We repeat the simulation of Fig. 1 but our estimates now assume a misspecified Ebola virus generation time distribution. This distribution has a mean that is 33% smaller than the one used to generate the epidemic data (which is from (Van Kerkhove et al., 2015)). Panel A provides estimates of instantaneous reproduction numbers, 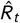, under the true and misspecified distributions (with 95% credible intervals) using EpiFilter (Parag, 2020). Panel B presents corresponding growth rate estimates (and 95% credible intervals), 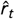, which are derived from the various 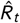 in A (Wallinga and Lipsitch, 2007).

## 4 DISCUSSION

Evaluating time-varying changes in pathogen transmissibility is an important challenge, allowing the impact of public health interventions to be assessed and providing indicators that can inform policymaking during epidemics. We have focussed on two key metrics for tracking transmissibility: the instantaneous reproduction number *R*_*t*_ (with estimate 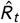) and the instantaneous growth rate *r*_*t*_ (with estimate 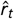). Both metrics provide key insights into the dynamics of epidemics as demonstrated by their use during the COVID-19 pandemic (Anderson et al., 2020; Abbott et al., 2020).

However, their relative merits and demerits have been increasingly debated. Recent work has suggested that the benefits of inferring 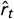 might have been underappreciated, and that this quantity may be particularly useful because of its apparent independence from modelling assumptions and its explicit consideration of the epidemic speed (i.e. it more naturally includes temporal information) (Pellis et al., 2020; Dushoff and Park, 2021).

Here, we have investigated and exposed the relationship between 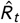 and 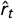. The relative informativeness of these two quantities during epidemics rests on the reliability of their smoothing and epidemiological assumptions. We found that both 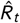 and 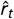 extract signals of changing pathogen transmission by smoothing noise from the incidence curve. As shown in Fig. 1, their key difference lies in the kernel (i.e., the set of weights in the SG filter of Eq. (2)) used for this smoothing. Specifically, computing 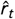 in a model-agnostic way corresponds to selecting an arbitrary kernel, whereas calculating 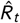 (and, correspondingly, model-based 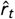 values) involves implicitly treating the generation time distribution as an epidemiological kernel (see Results and the right sides of Eq. (1) and Eq. (2)).

As a result, if the generation time distribution is estimated accurately and underlying assumptions about pathogen transmission hold, then not only are both measures closely related, with the commonly cited *R*_*t*_ = 1 threshold corresponding to an *r*_*t*_ = 0 threshold, but *R*_*t*_ is also theoretically more informative. This follows because 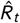 can be used to derive correct model-based 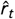 values, while also providing additional insights into the mechanism of transmission underlying the observed incidence (Yan and Chowell, 2019). In contrast, starting from the model-agnostic 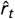, it does not seem possible to derive 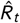 without epidemiological assumptions. Should the generation time distribution be misspecified as in Fig. 2, then 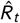 could be biased, and the model-agnostic 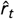 would be more informative.

When constructing 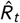, the generation time distribution is often approximated by the serial interval distribution. Misspecification of the generation time as described above might arise due to the often limited number of observed serial intervals used to estimate the serial interval distribution. Observed serial intervals are commonly obtained from household or contact tracing studies, where it is possible to identify source-recipient transmission pairs (Cowling et al., 2009; Li et al., 2020). However, as case numbers increase, identifying known source-recipient pairs becomes more challenging since there is less certainty about the source of a given transmitted infection and as the risk of infection from an unknown source in the community cannot be ignored.

Moreover, even if sufficient source-recipient pairs are reliably known, the generation time may still be misspecified. Non-pharmaceutical interventions and public health measures, such as case isolation after symptom onset, may curtail observed serial intervals (Ali et al., 2020) or increase the proportion of cases caused by pre-symptomatic transmission (Sun et al., 2021). In both scenarios it becomes difficult to reliably approximate the generation time distribution with the serial interval distribution, which is also now time-varying and may even have negative values. While recent approaches try to compensate for some of these issues (Ganyani et al., 2020) or allow the inclusion of up-to-date distributions (Thompson et al., 2019), accurately relating 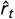 to 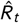 may not always be simple in practice.

Despite potential issues when obtaining 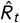, we have made clear that inferring the model-agnostic 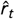 also requires assumptions related to smoothing of the incidence curve (or log incidence curve) and specification of the time interval over which to estimate a particular 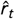. Furthermore, when case numbers are increasing, 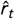 does not give an indication of the proportion of current transmissions that must be blocked to prevent an epidemic from continuing to grow. This proportion relative to *R*_0_ is known as the herd immunity threshold. This threshold is used to determine the vaccine coverage required in order to control transmission, accounting for vaccine effectiveness and any infection-acquired immunity (Hethcote, 2000; Thompson et al., 2020). On the other hand, it is 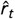 that naturally gives estimated doubling times (or halving times), which may be important for intervention planning purposes.

There are also a number of factors that limit the informativeness of both 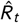 and 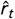. First, reporting errors and delays can lead to imprecise case counts, affecting summary statistics derived from incidence curves (Azmon et al., 2014). Second, both of the statistics discussed here relate to averages, but heterogeneous systems with superspreading individuals or events (Lloyd-Smith et al., 2005) require more than a measure of central tendency to be well understood. Inferring pathogen transmissibility and the potential impacts of interventions therefore often requires more complex modelling approaches. Last, it is not only 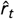 that requires time windows to be chosen for estimation. Values of 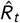 are often calculated over shifting time windows. Short windows may lead to fluctuating 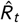 values that potentially reflect randomness in contacts between hosts rather than variations in transmissibility, while long windows may blur detection of key variations (Cori et al., 2013; Parag and Donnelly, 2020).

The above problems relate to fundamental bias-variance tradeoffs in the inference of *r*_*t*_ and *R*_*t*_ and emphasise that neither measure should be used naively. As highlighted in (Lloyd, 2009) and illustrated in Fig. 2, sensitivity analyses of the structure of the epidemiological model or statistical procedure used are crucial for drawing reliable inferences from noisy data. It should also be noted that even if these problems do not exist, other contextual information is still often required to obtain a full picture of an ongoing epidemic. For example, while *R*_*t*_ = 1 or equivalently *r*_*t*_ = 0 may indicate a stable epidemic, the policy response may be very different depending on whether incidence is high or low. The first of these scenarios may not be acceptable to policymakers, as it involves large numbers of infections in the near future. Both *R*_*t*_ and *r*_*t*_ only provide information about the changes in state of an epidemic.

Nonetheless, despite some of the challenges in estimation and the need for contextual information, we contend that both 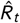 and 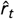 are valuable. Estimates of *R*_*t*_ (widely referred to as the “R number”) are particularly useful as an intuitive measure for public communication, allowing the effects of current interventions to be assessed and communicated straightforwardly. However, estimates of *r*_*t*_, expressed as doubling times, are great for expressing the speed at which cases are increasing. Given the risks of depending on either *R*_*t*_ or *r*_*t*_ that we have explored in this paper and the complementary roles they can play in raising public awareness, we support current efforts to generate estimates of both summary statistics. These quantities in combination and together with contextual measures such as current incidence or prevalence, allow epidemic dynamics to be understood more clearly and completely.

## Data Availability

This paper generates no data.

